# Comparing the risks of diabetes, psychological distress, and radiation-induced cancer exposure after the Fukushima disaster using the loss of happy life expectancy

**DOI:** 10.1101/2023.05.01.23289327

**Authors:** Michio Murakami, Akihiko Ozaki, Kyoko Ono, Shuhei Nomura, Yoshitake Takebayashi, Masaharu Tsubokura

## Abstract

After disasters, direct deaths and secondary health effects, such as diabetes and psychological distress, can occur. It is critical to compare the magnitudes of these risks to promote effective public health measures. In this study, we used the loss of happy life expectancy (LHpLE) to compare the risks associated with diabetes, psychological distress, and radiation-induced cancer after the 2011 Fukushima disaster. Two questionnaire surveys were conducted on people affected by the disaster to investigate the potential associations between diabetes and psychological distress, as well as breast cancer and reduced emotional happiness, with 680 and 582 participants, respectively. Additionally, we calculated the LHpLE owing to these risks. Although no significant reduction was found in emotional happiness due to diabetes or breast cancer, a significant reduction occurred due to psychological distress (0.265 and 0.476 for males and females, respectively). In the population aged 40–74 years, the LHpLE due to radiation-induced cancer, diabetes, and psychological distress were 0.0013, 0.14, and 0.21 years, respectively. This suggests that the association of LHpLE with diabetes and psychological distress was over two orders of magnitude greater than that associated with radiation-induced cancer. Within 7 years following the disaster, LHpLE due to diabetes increased, highlighting that diabetes is an ongoing issue. Therefore, this novel indicator of the LHpLE can provide a foundation for promoting effective public health measures following disasters.

## 1. Introduction

Disasters cause direct deaths and have secondary physical and mental health effects, which lead to associated indirect deaths. These effects are exacerbated by regulations and life changes implemented in response to disasters. For example, Hurricane Katrina in the United States in 2005 resulted in an increase in lifestyle-related diseases, such as diabetes, following prolonged evacuation [1]. Additionally, a systematic review found that those affected by natural disasters experience increased psychological distress and psychiatric disorder [2]. The 1986 Chornobyl nuclear power plant accident also resulted in worsened mental health among those affected, which was associated with radiation anxiety [3]. Even after the outbreak of coronavirus disease 2019 (COVID-19), an increase in diabetes and psychiatric disorders has been reported [4, 5]. Comprehensive health promotion policies that address both direct and secondary health effects are becoming increasingly important in the aftermath of disasters [6].

The 2011 Great East Japan Earthquake and Fukushima Daiichi Nuclear Power Station accident (hereafter referred to as the Fukushima disaster) had various secondary health effects. According to the United Nations Scientific Committee on the Effects of Atomic Radiation (UNSCEAR), no statistically identifiable increase was found in radiation-induced cancer after the Fukushima disaster due to the immediate implementation of radiation dose reduction measures [7]. However, the Fukushima disaster led to over 140,000 mandatory evacuees [8], and secondary physical and mental health effects, such as the increased prevalence of diabetes and psychological distress, have been consistently reported in several municipalities [4, 9-12]. Therefore, it is essential to compare the magnitudes of risk using consistent indicators to effectively address these diverse health effects. Risk indicators, such as mortality [13], loss of life expectancy (LEE) [14], and disability-adjusted life years (DALYs) [15], have been proposed to compare the magnitude of various risk events and facilitate effective policymaking. For example, Murakami et al. employed LEE post-Fukushima disaster to compare evacuation from nursing homes and increased diabetes prevalence with the risk of radiation-induced cancer [16, 17]. Based on the premise that risk events encompass not only increased mortality but also diminished well-being, the loss of happy life expectancy (LHpLE; shortened length of lifespan during which a person experiences subjective emotional happiness) indicator was proposed to compare increased psychological distress among those affected by the Fukushima disaster with the risk of radiation-induced cancer [18]. The LHpLE risk indicator is computed by integrating a life table and an emotional happiness questionnaire, extending the quality-of-life-weighted risk indicators, such as DALYs. This promising indicator allows for a comparison of the magnitude of different types of risks, such as cancer and psychological distress, while also reflecting the value system of disaster-affected populations regarding improved well-being [19]. Nonetheless, the LHpLE risk indicator has not been applied to other secondary health effects, including the escalation of diabetes prevalence. Moreover, the risk of radiation-induced cancer has not been thoroughly assessed regarding its impact on emotional happiness except in a limited number of studies (preprint; [20]). Given the recent accumulated knowledge on long-term radiation exposure, diabetes prevalence, and psychological distress after the Fukushima disaster, comparing the magnitude of risk associated with these exposure factors using the LHpLE indicator could have important implications for future reconstruction in Fukushima and subsequent post-disaster policymaking.

Therefore, in this study, we first investigated whether diabetes and psychological distress reduced emotional happiness through a survey of residents in former evacuation-order areas affected by the Fukushima disaster. Second, we examined whether cancer reduces emotional happiness through a survey of patients with breast cancer (which has the highest incidence rate among Japanese females) and patients without breast cancer (such as those who underwent health checkups and outpatient care) in a hospital in Fukushima Prefecture. Third, we quantitatively compared the risks of diabetes, psychological distress, and radiation-induced cancer after the Fukushima disaster using the LHpLE indicator.

## 2. Methods

### 2.1. Ethics

The Ethics Committees of Fukushima Medical University (General 29199) and the Jyoban Hospital of Tokiwa Foundation (JHTF-2021-006) approved this study. Participants’ consent was obtained from a questionnaire survey.

### 2.2. Two surveys

#### Emotional happiness and psychological distress and diabetes

We analyzed whether psychological distress or diabetes caused a decrease in emotional happiness among residents aged 20 years and older in former evacuation order areas using a mailing questionnaire (conducted in January 2018, covering 2000 participants, with a total of 680 valid responses). The details of the questionnaire survey have been reported in a previous study [21].

#### Emotional happiness and breast cancer

To test whether there was a reduction in emotional happiness due to cancer, we administered a questionnaire from November 2021 to January 2022 to patients with and without breast cancer aged 20 years or older who were outpatients of the Department of Breast and Thyroid Surgery, Jyoban Hospital, Fukushima. Information on whether the patient had breast cancer was based on medical records. Of the 724 patients distributed, 562 with medical record information who provided consent and responded in person were included in the study. All patients who responded according to sex were female.

Both surveys asked participants about emotional happiness using the following question: “Did you experience a feeling of enjoyment yesterday? [Yes or No]” [18, 21, 22]. In a survey regarding the reduction in emotional happiness due to breast cancer, we also asked about the following covariates in the association between breast cancer and emotional happiness, as previously conducted in another study[18]: subjective feelings of health, psychological distress, job, marital status, children, grandchildren, educational background, joblessness within the household, annual household income, smoking habits, and age. Psychological distress was measured using the Kessler-6 (K6) scale, with a cutoff of 12/13 [23]. Residents (Hamadori and others) were also surveyed. Additionally, given that this survey was implemented after the outbreak of COVID-19, we inquired about impediments to life due to the COVID-19 pandemic.

### 2.3. LHpLE

Details on the calculation of happy life expectancy (HpLE) and LHpLE are described in a previous study [18]: HpLE is calculated from the life table by sex (*s*) and the percentage of emotional happiness (*Hp_x,s_*) by sex and age (*x*) groups), according to equations 1 and 2.

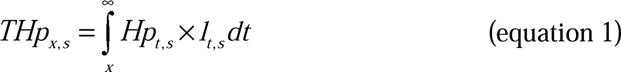

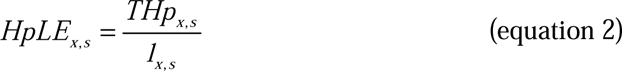

where *l_x,s_*is the number of people living at the start of an interval at age *x* ( 100,000 born alive).

For the life table and *Hp_x,s_*, we used the respective values for Japan as a whole according to previous reports [18, 24]. For *Hp_x,s_*, we used values for males and females in their 20s, 30s, 40s, 50s, and 60s. For those under the age of 20 and those aged 70 years or older, we used the values for those in their 20s and 60s. Additionally, LHpLE*_x,s_* for a given sex *s* and age *x* is calculated as shown in equation 3 by computing HpLE*_x,s_*’ considering the decrease in emotional happiness and the increase in mortality due to risk events.

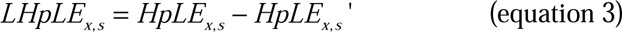

### 2.4. Statistical analysis: decrease in emotional happiness

#### Emotional happiness and psychological distress and diabetes

To analyze the reduction in emotional happiness due to psychological distress and diabetes, we conducted sex-specific propensity score matching using variables that were significantly associated with positive emotions (including emotional happiness and similar items) as covariates, as in a previous study [21]. For psychological distress, we used the following covariates for males: subjective feelings of health, dyslipidemia history, marital status, joblessness within the household, and smoking habits. For females, we used subjective feelings of health, age, and evacuation status as covariates. Psychological distress was also included as a covariate of diabetes. Propensity score matching was performed using a 1:1 nearest-neighbor matching and a ±0.1 caliper without replacement, as conducted in a previous study [18]. We obtained 34 (mean caliper: 0.0012) and 21 (mean caliper: 0.0000) matched pairs for males and females with psychological distress, respectively, and 61 (mean caliper: 0.0027) and 24 (mean caliper: 0.0000) matched pairs for males and females with diabetes, respectively. McNemar’s tests were conducted separately for males and females to examine the differences in emotional happiness between those with and without psychological distress or diabetes after matching.

#### Emotional happiness and breast cancer

The analysis of the decline in emotional happiness due to breast cancer was conducted as follows: first, we examined the associations of emotional happiness with individual attributes, including subjective feelings of health, psychological distress, impediments to life due to the COVID-19 pandemic, residence, job, marital status, children, grandchildren, educational background, joblessness within the household, annual household income, smoking habit, age, or breast cancer using the chi-square test, Fisher’s exact test, or Fisher-Freeman-Halton’s exact method. As subjective feelings of health, marital status, presence of a child, presence of an unemployed person within the household, and psychological distress were significantly associated with emotional happiness (P < 0.05), we included these items as covariates in propensity score matching. Subsequently, propensity score matching and McNemar’s tests were conducted, as described above. Propensity score matching yielded 161 matched pairs (mean calipers: 0.0002). All analyses were performed using the IBM SPSS Statistics 24 or 28 or R statistical software [25, 26].

### 2.5 Calculation of LHpLE after the Fukushima disaster

Because the results of the above analyses showed a significant decrease in emotional happiness due to psychological distress for both males and females (P < 0.05) (see “**3.1. Decline in emotional happiness due to psychological distress, diabetes, and breast cancer”**), we included the decrease in emotional happiness Δ*Hp* (−0.265 and −0.476 for males and females, respectively) when calculating the LHpLE for psychological distress. Since no significant decrease was found in emotional happiness due to diabetes, the decrease in emotional happiness due to diabetes was not considered. Similarly, no significant decrease was observed in emotional happiness due to breast cancer. Additionally, a previous study (preprint; [20]) reported that patients with various types of cancer, regardless of sex, exhibit a decrease in emotional happiness in Japan as a whole. Therefore, the decline in emotional happiness due to breast cancer was not considered in the LHpLE calculations. We did not consider the increase in mortality for psychological distress but considered it for diabetes and radiation-induced cancer after the Fukushima disaster.

For psychological distress and radiation-induced cancer, we calculated the LHpLE for males and females aged 20, 30, 40, 50, 60, 70, and 80 years and sex- and age-adjusted LHpLE for those aged 20 and older and 40 to 74 years using the 2015 model population [27]. For diabetes, we calculated LHpLE for 40-, 50-, 60-, and 70-year-old males and females and age-adjusted LHpLE for 40- to 74-year-olds. We also calculated the sex- and age-adjusted LHpLE for patients aged 20 years and older and for those aged 40–74 years and determined the ratio of LHpLE for these risks to LHpLE.

#### Psychological distress

The increase in the prevalence of psychological distress after the Fukushima disaster (*AP*) was calculated by subtracting the values at the normal time (0.03) from those for residents in the evacuation order areas for 8 years after the disaster (0.146, 0.117, 0.097, 0.077, 0.071, 0.068, 0.064, and 0.057 in order from years 1 to 8) [28]. *AP,* since the 8th year after the disaster, was assumed to decay exponentially. *AP* was assumed not to differ by sex or age. Emotional happiness with risk at age *x* (*Hp_x_’*) was calculated using equation 4 as follows:

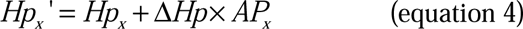

#### Diabetes

For diabetes, we used the results of individuals aged 40–74 years who underwent health checkups in Minamisoma City as a proxy for those affected by the Fukushima disaster [9]. An increase in diabetes has been consistently reported in municipalities with evacuation order areas other than Minamisoma City [10, 11]. We calculated the additional prevalence for 7 years after the disaster from the pre-disaster prevalence for non-returnees (3.48%) and the 1-year odds ratio (1.18) [9]. Specifically, we assumed that age-adjusted prevalence would not change in the absence of a disaster and that an increase in prevalence in 1 year would be considered as the incidence of diabetes occurring in that year. We calculated the incidence rates for the first to seventh years after the disaster, assuming that diabetes occurred in males and females aged 40, 50, 60, and 70 years at the time of the disaster at the same rate; the increase in prevalence in the eighth year after the disaster was not considered. The increase in diabetes-related mortality was based on the results of a previous study [29]. We used the hazard ratios (*HR*) in a cohort study on increased all-cause mortality due to a history of diabetes in Japan (17.8 years of central follow-up, totaling 99,584 participants) and the Japanese Life Table [24] as the basis for the calculation. *HRs* were 1.20 and 1.59 for males and 1.45 and 2.00 for females within 15 years and >15 years of onset, respectively. The increase in mortality due to diabetes *A(x,s)* was calculated as follows:

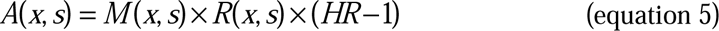

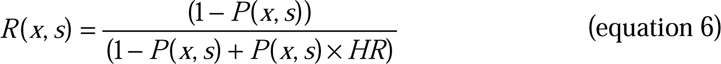

where *M(x,s)* is the all-cause mortality rate, *R(x,s)* is the ratio of all-cause mortality among those without diabetes to the total, and *P(x,s)* is the prevalence of diabetes. We calculated *R(x,s)* from the *P(x,s)* in 2012 in Japan [30] and the *HR* (1.59 and 2.00 for males and females, respectively).

LHpLE due to diabetes in the entire target population was calculated by multiplying the LHpLE among patients with diabetes by the proportion of these patients.

#### Radiation-induced mortality

Mortality calculation was slightly modified from that of previous studies [17, 18]. The modifications included updating the dose. Specifically, based on the UNSCEAR report [7], an average value for all evacuees was calculated from the population of residents in each municipality in the evacuation order areas and evacuation scenario, and the first and lifetime exposure (up to age 80) dose of a 20-year-old adult (0.96 mSv in the first year and 14.5 mSv lifetime exposure). Doses were assigned for the second and subsequent years to account for the physical decay of radionuclides [31]. The effective dose was considered equal to the organ doses for all solid cancers and leukemia (i.e., 1 Sv = 1 Gy). For the dose-response equations, we used excess relative risk (ERR) model equations for all solid cancers [32] and leukemia [33, 34] (*ERR_solid_* and *ERR_leukemia_*) based on the Life Span Study cohort among Atomic Bomb survivors in Hiroshima and Nagasaki. Furthermore, we calculated the theoretical increase in mortality by sex and age using a linear-quadratic dose-response model, which showed a better fit than the linear non-threshold model under low doses [32]. The following equations were used to calculate *ERR_solid_* and *ERR_leukemia_*:

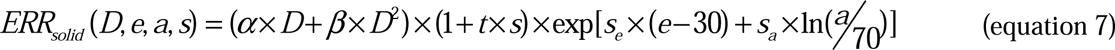

where *D* is the organ dose (Sv), *e* is the age at exposure, *a* is the age attained, *s* is sex (−1 for men, +1 for women), α = 0.22, β = 0.18, *t* = 0.29, *s_e_* = −0.034, and *s_a_* = −0.89.

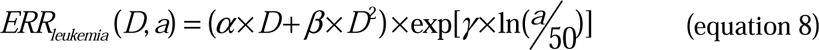

where α = 1.612, β = 1.551, and γ = −1.634.

The minimum latency period was 5 and 2 years for all solid cancers and leukemia, respectively. Mortality rates for all solid cancers and leukemia were based on statistical data from age- and sex-stratified all-cause mortalities in Japan [35].

## 3. Results

### 3.1. Decline in emotional happiness due to psychological distress, diabetes, and breast cancer

Table 1 shows the relationships between individual attributes or breast cancer, and emotional happiness. Subjective feelings of health, psychological distress, marital status, the presence of children, and the presence of joblessness within the household were significantly associated with emotional happiness (P < 0.05). However, no significant association was found between breast cancer and emotional happiness (P = 0.472).

**Table 1.** Associations between emotional happiness and individual attributes.

| Items |  | Emotional happiness: N (%) |  | P |
| --- | --- | --- | --- | --- |
|  |  | No | Yes |  |
| Subjective feelings of health | Low | 86 (27%) | 228 (73%) | <0.001 |
|  | High | 23 (13%) | 148 (87%) |  |
| Psychological distress | Low | 94 (21%) | 360 (79%) | <0.001 |
|  | High | 21 (49%) | 22 (51%) |  |
| Impediments to life due to COVID-19 pandemic | Not at all | 13 (22%) | 46 (78%) | 0.590 |
|  | Not so much | 34 (27%) | 90 (73%) |  |
|  | To some degree | 53 (23%) | 180 (77%) |  |
|  | Very much | 16 (20%) | 66 (80%) |  |
| Residence | Hama-dori | 112 (23%) | 380 (77%) | 0.054 |
|  | Other | 4 (57%) | 3 (43%) |  |
| Job | Company employees etc. | 37 (21%) | 137 (79%) | 0.116 |
|  | Self-employed | 1 (6%) | 17 (94%) |  |
|  | Other | 76 (26%) | 222 (74%) |  |
| Marital status | Unmarried | 19 (38%) | 31 (62%) | 0.004 |
|  | Married, living together | 67 (20%) | 269 (80%) |  |
|  | Married, living separately | 8 (50%) | 8 (50%) |  |
|  | Divorced | 15 (25%) | 44 (75%) |  |
|  | Widowed | 6 (17%) | 30 (83%) |  |
| Children | Presence | 85 (21%) | 322 (79%) | 0.026 |
|  | Absence | 29 (32%) | 61 (68%) |  |
| Grandchildren | Presence | 31 (23%) | 102 (77%) | 1.000 |
|  | Absence | 74 (23%) | 246 (77%) |  |
| Educational background | Junior or high-school graduate | 64 (26%) | 181 (74%) | 0.057 |
|  | University etc. graduate | 46 (19%) | 196 (81%) |  |
|  | Do not answer | 2 (50%) | 2 (50%) |  |
| Joblessness within the household | Presence | 14 (47%) | 16 (53%) | 0.004 |
|  | Absence | 95 (21%) | 357 (79%) |  |
|  | Do not answer | 0 (0%) | 2 (100%) |  |
| Annual household income | < 3 million Japanese Yen | 39 (27%) | 105 (73%) | 0.295 |
|  | 3–6 million Japanese Yen | 29 (22%) | 104 (78%) |  |
|  | ≥ 6 million Japanese Yen | 26 (18%) | 120 (82%) |  |
|  | Do not answer | 16 (24%) | 50 (76%) |  |
| Smoking habit | Do not smoke | 98 (22%) | 352 (78%) | 0.348 |
|  | Do smoke | 13 (29%) | 32 (71%) |  |
| Age | 20–30s | 7 (12%) | 53 (88%) | 0.117 |
|  | 40s | 33 (26%) | 94 (74%) |  |
|  | 50s | 34 (29%) | 85 (71%) |  |
|  | 60s | 15 (21%) | 58 (79%) |  |
|  | 70s or more | 15 (21%) | 55 (79%) |  |
| Breast cancer | Absence | 82 (24%) | 254 (76%) | 0.472 |
|  | Presence | 47 (22%) | 171 (78%) |  |

Figure 1 shows the differences in emotional happiness between the presence and absence of diseases (i.e., psychological distress, diabetes, and breast cancer) after propensity score matching. Emotional happiness was 0.265 and 0.476 lower for males and females with psychological distress, respectively (P < 0.05). However, no significant differences were observed in diabetes (in both males and females) or breast cancer (P > 0.200).

**Figure 1.**
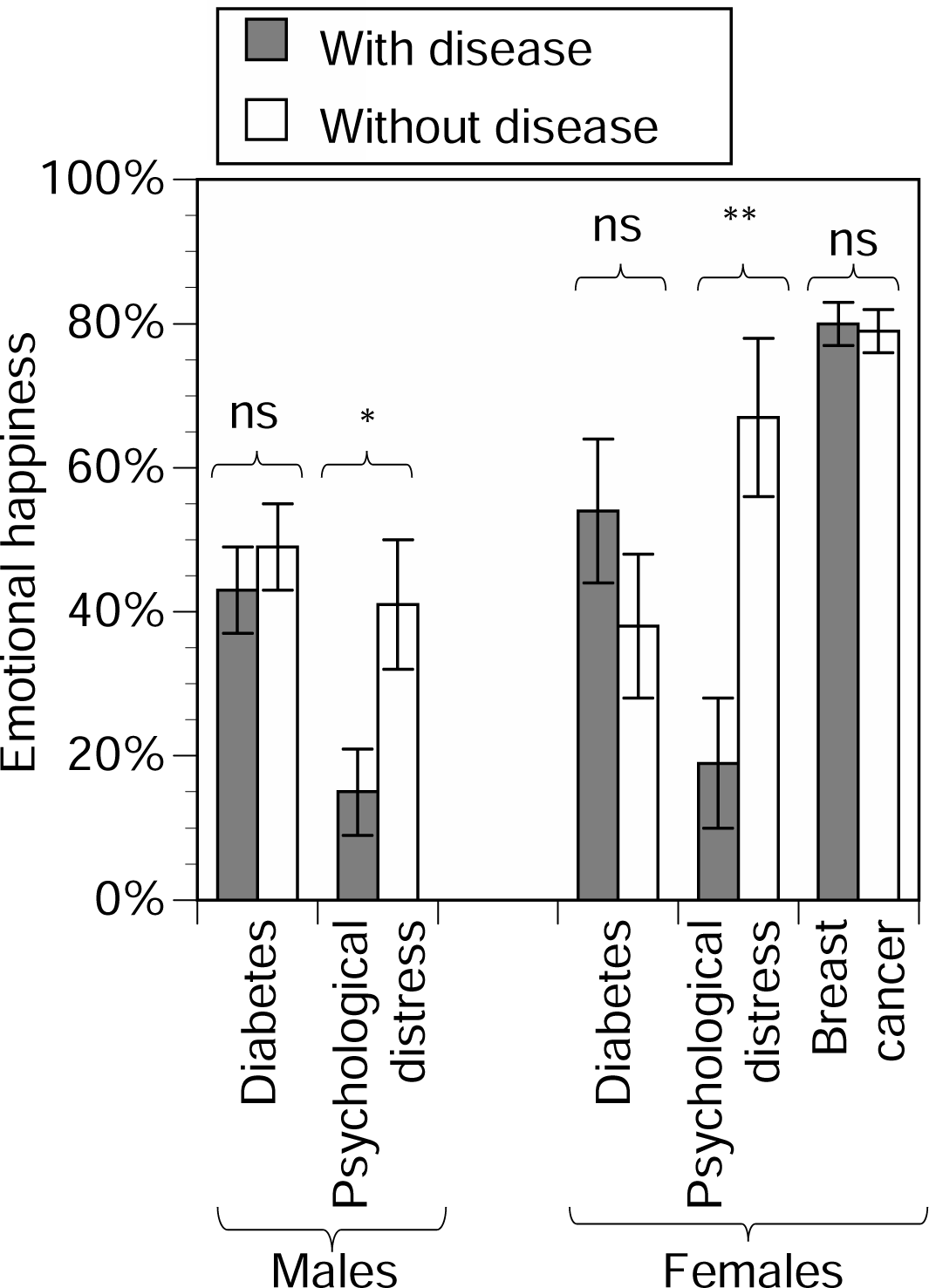
Comparison in emotional happiness between the presence and absence of diseases. Error bars represent standard error. * P < 0.05, ** P < 0.01, ns: not significant. Matching regarding psychological distress among males involved adjustment for subjective feelings of health, dyslipidemia history, marital status, presence of a jobless person within the household, and smoking habit as covariates. Matching regarding psychological distress among females involved adjusting for subjective feelings of health, age, and evacuation status as covariates. For diabetes, psychological distress was added as a covariate. Matching regarding breast cancer among females involved adjusting for subjective feelings of health, marital status, presence of children, the presence of a jobless person within the household, and psychological distress as covariates.

### 3.2. LHpLE for diabetes, psychological distress, and radiation-induced cancer after the Fukushima disaster

Figure 2 shows the comparison of LHpLEs for diabetes, psychological distress, and radiation-induced cancer following the Fukushima disaster. The LHpLE due to diabetes (for the 7-year post-disaster period only) was 0.14 years in the 40–74 age group. Of these, LHpLE due to increased diabetes in the first, second to fourth, and fifth to seventh years was 0.012, 0.049, and 0.075 years, respectively. The LHpLE due to psychological distress was 0.20 and 0.21 years for those aged 20 years and older and 40–74 years, respectively. The LHpLE associated with increased psychological distress in the first, second to fourth, fifth to seventh, and eighth years for those aged 20 years and older were 0.043, 0.073, 0.040, and 0.046, respectively. Similarly, for the 40–74 year age group, the LHpLE was 0.043, 0.074, 0.041, and 0.048 years, respectively.

**Figure 2.**
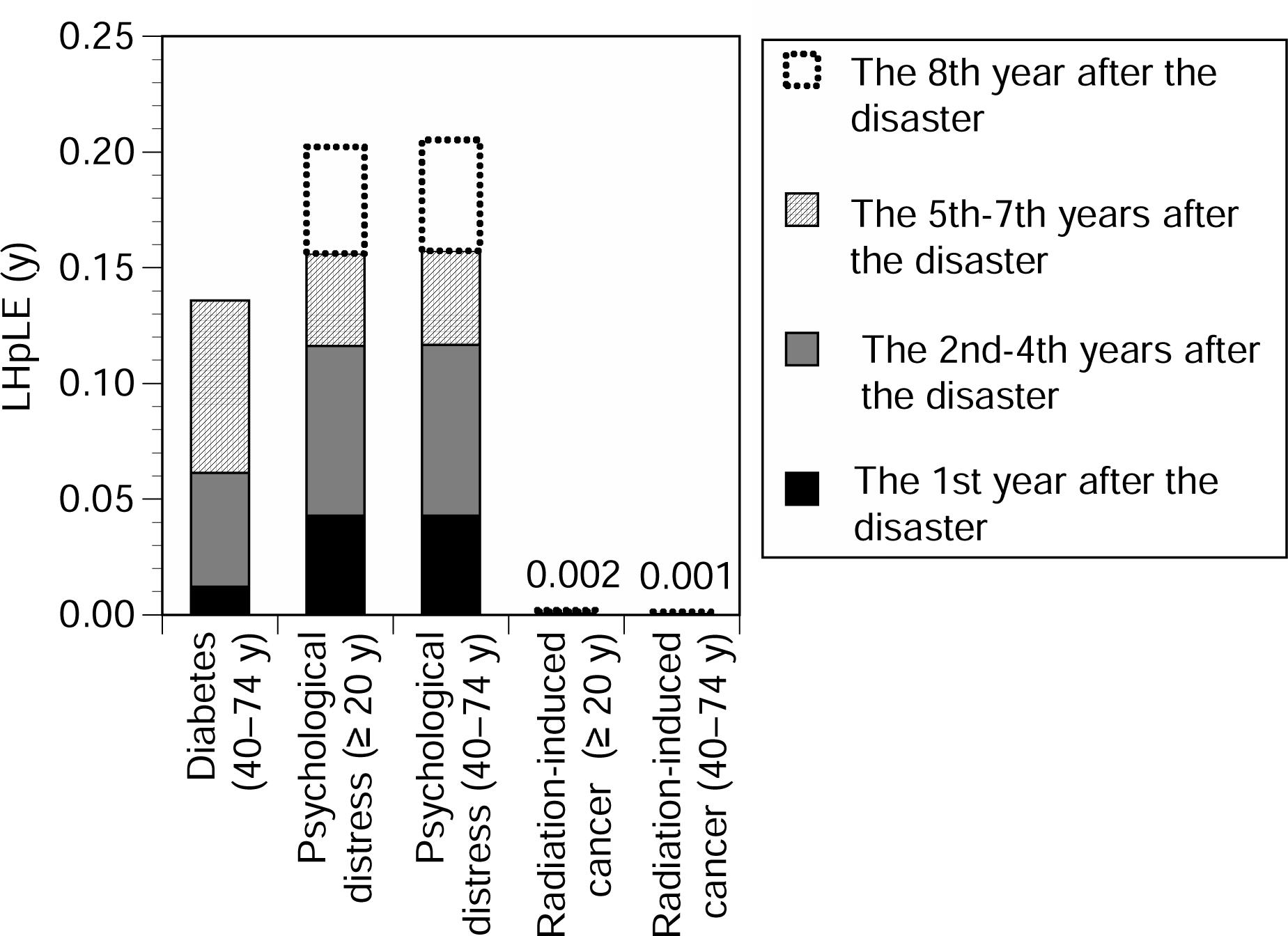
Comparisons of loss of happy life expectancy (LHpLE) among diabetes, psychological distress, and radiation-induced cancer. LHpLE was categorized into four time periods depending on when the risk occurred. Diabetes indicates the time of new-onset; psychological distress indicates its prevalence; and radiation-induced cancer indicates the time of radiation exposure.

The LHpLE due to radiation-induced cancer after the Fukushima disaster was 0.0020 years for affected people aged 20 years and older and 0.0013 years for those aged 40–74 years. The LHpLE due to radiation-induced cancer in the first, second to fourth, fifth to seventh, and eighth years for those aged 20 years and older were 0.00026, 0.00062, 0.00032, and 0.00081 years, respectively. Similarly, for the 40–74 year age group, they were 0.00019, 0.00044, 0.00022, and 0.00044 years, respectively.

The ratio of LHpLE due to diabetes to HpLE was 6.9 × 10^-3^ for ages 40–74 years, while that due to psychological distress was 9.2 × 10^-3^ and 1.0 × 10^-2^ for those aged 20 years and older and 40–74 years, respectively. The ratios of LHpLE due to radiation-induced cancer to HpLE were 9.2 × 10^-5^ and 6.6 × 10^-5^ for those aged 20 years and older and 40–74 years, respectively.

## 4. Discussion

In this study, we quantitatively compared three different types of risks ( diabetes, psychological distress, and radiation-induced cancer) after the Fukushima disaster using the LHpLE indicator. First, our findings confirmed that no significant decrease in emotional happiness was linked to breast cancer or diabetes, whereas a significant decrease was observed in psychological distress (0.265 and 0.476 for males and or females, respectively). This is consistent with previous research showing no significant decline in emotional happiness among patients with cancer in Japan (preprint [20]). Our analysis, which employed propensity score matching, revealed no significant association between diabetes and positive emotion [21], aligning with the results of the same questionnaire surveys used in this study. In contrast, it has been reported that in Japan, emotional happiness decreased by 0.19 and 0.22 for males and females, respectively, due to psychological distress [18]. However, this study, which was conducted among former evacuation-order area residents, demonstrated a larger decline. A possible explanation could be the difference in the severity of psychological distress between the Fukushima disaster-affected and Japanese populations. In this study, a score of 13 or higher on the K6 was considered psychological distress, which is consistent with the clinical diagnosis [23]. It is possible that the psychological distress among the disaster-affected population might be more severe than in Japan as a whole. Our results highlight the importance of calculating the decline in emotional happiness based on the participants’ responses to questionnaires when assessing LHpLE more accurately.

Among the patients with diabetes, psychological distress, and radiation-induced cancer after the Fukushima disaster, LHpLE due to diabetes and psychological distress was more than two orders of magnitude higher than that due to radiation-induced cancer. Particularly, the ratios of LHpLE due to diabetes and psychological distress to HpLE, ranging from 6.9 × 10^-3^ to 1.0 × 10^-2^, demonstrated that diabetes and psychological distress after the disaster were not negligible. The finding that diabetes has the same magnitude of risk as psychological distress after the Fukushima disaster is important. The Chornobyl nuclear power plant accident had the greatest impact on mental health [36]. The secondary health effects of a nuclear disaster are not limited to mental health but also indicate that physical risks, such as diabetes, can be serious. In examining temporal changes until the seventh year post-disaster, LHpLE due to psychological distress was attenuated from the post-disaster period onwards, whereas LHpLE due to diabetes increased over time. These results have significant implications for public health policy-making following complex disasters. First, providing comprehensive health support in the aftermath of a nuclear disaster is critical, addressing radiation exposure measures and secondary health effects, such as psychological distress and diabetes. Because radiation exposure levels depend on the severity of the disaster, this does not imply that in any nuclear disaster, mitigating psychological distress and secondary health effects, such as diabetes, will be prioritized over radiation exposure. Nevertheless, the risks of diabetes and psychological distress remain high, necessitating a comprehensive and balanced combination of public health measures that address both radiation protection and the secondary health effects in nuclear disaster scenarios.

Second, this study provides fundamental insights into a timely approach to reconstruction assistance following the Fukushima disaster. Specifically, the observation that diabetes remains a significant risk factor for several years into the chronic post-disaster phase highlights the importance of sustained support for recovery. Although this study did not assess the incidence of diabetes after the eighth year post-disaster owing to uncertainties, the possibility of a future increase cannot be ruled out. Therefore, long-term monitoring and prevention of the onset of diabetes are necessary.

Furthermore, a previous study estimated the HpLE gain resulting from returning to be 0.8 years for 40-year-old females [37], which was larger than the LHpLE due to diabetes and psychological distress. This implies the importance of maintaining or improving daily well-being while addressing radiation exposure, diabetes, and psychological distress post-disaster. Among the Fukushima disaster-affected population, factors such as household unemployment and living apart from partners have been reported to reduce well-being [21]. Public health measures related to radiation protection and physical and mental health should run parallel to factors that support well-being, such as employment and family life.

This study had some sources of uncertainies. Overall, the calculations were based on the assumption that the risk of radiation-induced cancer could be overestimated, while the risks of diabetes and psychological distress could be underestimated [17, 18]. Specifically, the models for calculating mortality were based on epidemiological studies of Atomic Bomb survivors in Hiroshima and Nagasaki, Japan. Using models with exposure to low doses provided conservative assessments (i.e., overestimated values). Furthermore, these calculations do not consider future improvements in cancer treatment. Although this study found that emotional happiness was not reduced by cancer among patients with breast cancer who were affected by the Fukushima nuclear disaster, it was impossible to examine this for other types of cancer. However, as described above, no decrease was observed in emotional happiness among cancer patients throughout Japan (preprint, [20]).

The assumption that diabetes would exhibit no change in age-adjusted prevalence if there had been no disaster aligns with the observation that no increase in age-adjusted diabetes prevalence has been observed in Japan as a whole [38]. The literature used to calculate diabetes prevalence might have underestimated it because it was based on glycated hemoglobin levels rather than disease history, which was used to estimate *HR* in a previous cohort study [29]. The incidence of diabetes may have increased after the eighth year post-disaster; however, this study excluded its contribution.

Additionally, the cohort used to calculate the increase in mortality excluded individuals with cardiovascular disease, chronic liver disease, kidney disease, or any type of cancer at baseline [29]. Because people with such diseases are more likely to have diabetes, the *HR* used in this study may have been underestimated.

In this study, we did not consider a possible increase in mortality rates due to psychological distress in the study [39]. The reduction in emotional happiness due to psychological distress might have been adjusted for covariates within the items measured rather than fully adjusted for. Although the risks of diabetes and psychological distress may occur synergistically, each risk factor was treated independently in this study. Therefore, the risk of having both of these factors has not yet been evaluated. To calculate the LHpLE, we substituted values for emotional happiness in the 20s and 60s for those under 20 and 70 years or older, respectively. A previous study showed that differences due to this substitution were negligible [18]. Since this study did not consider differences by age or sex in the incidence of diabetes or the prevalence of psychological distress, the LHpLE was calculated as the average for males and females aged 40–74 years or 20 years and older. Thus, more detailed data are required to elucidate the differences in risk according to age and sex.

Despite these uncertainties, our conclusion remains unaffected: LHpLE due to diabetes and psychological distress were greater than that due to radiation-induced cancer.

## 5. Conclusions

In this study, we examined the potential decrease in emotional happiness due to diabetes, psychological distress, and breast cancer using questionnaire surveys administered to people affected by the Fukushima disaster. Furthermore, we quantitatively compared the risks of diabetes, psychological distress, and radiation-induced cancer among the affected population using the LHpLE indicator. The key findings of this study are summarized as follows:

- Although no decline in emotional happiness was observed among those affected by the Fukushima disaster because of diabetes or breast cancer, a significant decrease in emotional happiness associated with psychological distress was detected.
- The LHpLE allows for a comparison of various types of risks, such as cancer, diabetes, and psychological distress. During the Fukushima disaster, the incidence of LHpLE due to diabetes and psychological distress was more than two orders of magnitude greater than that due to radiation-induced cancer. The ratios of LHpLE due to diabetes and psychological distress to HpLE ranged from 6.9 × 10^-3^ to 1.0 × 10^-2^, highlighting that these risks were not negligible.
- LHpLE due to diabetes increased over time during the seven years following the disaster, suggesting that addressing diabetes remains an important issue.
- The comparisons of different types of risks using the LHpLE can provide fundamental information for formulating public health measures that comprehensively balance radiation protection and the secondary health effects of nuclear disasters.

## Notes

Preliminary results were presented at the 2022 Annual Conference of the Society of Environmental Science in Japan [40].

## Author contribution

Michio Murakami: Conceptualization; Data curation; Methodology; Formal analysis; Visualization; Project administration; Writing original draft.

Akihiko Ozaki: Conceptualization; Data curation; Writing –review & editing.

Kyoko Ono: Conceptualization; Writing –review & editing.

Shuhei Nomura: Conceptualization; Writing –review & editing.

Yoshitake Takebayashi: Conceptualization; Writing –review & editing.

Masaharu Tsubokura: Conceptualization; Writing –review & editing.

## Data Availability

All data produced in the present study are available upon reasonable request to the authors.

## Acknowledgements

We would like to thank Editage (www.editage.com) for English language editing. This work was supported by JSPS KAKENHI Grant Number JP17K20069, JP20H04354 and “The Nippon Foundation - Osaka University Project for Infectious Disease Prevention.” The external funders have no role in study design, data collection and analysis, decision to publish, or preparation of the manuscript.

